# Universal Screening with a Glucose Challenge Test to Predict Gestational Diabetes Mellitus in a Diverse Population: A Retrospective Cohort Study

**DOI:** 10.64898/2026.10.06.26364834

**Authors:** Xin Yi Jolynne Tan, Yu Xuan Huang, Mark Pandrich, Gabriele Kerr, Jennifer Thornton, Richard A. Powell, Jonathan Valabhji, Ana Neves, Nishel Shah, Mark Johnson, Natasha Singh

## Abstract

**Introduction:** Gestational diabetes mellitus (GDM) is associated with significant maternal and neonatal morbidity. Screening strategies vary internationally and the diagnostic performance of the 50g glucose challenge test (50g GCT) as a universal screening tool remains uncertain across diverse populations.

**Research design and methods:** This retrospective cohort study included 18,213 singleton pregnancies at a London tertiary hospital between 2010 and 2023. Women routinely underwent the universal two-step GDM screening pathway: a 50g GCT at 24–32 weeks’ gestation, followed by a diagnostic 75-g oral glucose tolerance test if the 50g GCT result was ≥7.8 mmol/L. We evaluated the use of universal two-step screening in detecting GDM, how its prevalence varies with maternal characteristics, including ethnicity, body mass index, age, and in those who would not have been included in risk factor-indicated screening, and the association between the 50g GCT and adverse outcomes.

**Results:** 2,015 women had an abnormal GCT, and 406 (20.1%) had an abnormal 75g OGTT, with GDM diagnosed at 24-28 weeks. We also found 87 women, despite a normal GCT and/or normal OGTT, developed late-onset GDM. The overall prevalence of GDM was 4.4% (810/18,213). Universal screening will therefore identify a significant number of GDM cases, notably including those in a low-risk cohort who would not have been screened using a risk factor-based approach. When we examined glucose as a continuous variable, there was an association between increasing 50g GCT values and adverse maternal and neonatal outcomes.

**Conclusion:** Universal screening with a GCT is a feasible and safe approach and addresses a significant health disparity gap, in that women without risk factors are screened and may be diagnosed with GDM. Given the long-term cardiometabolic implications of GDM, pregnancy diagnosis is important to allow for longer-term targeting of cardiometabolic risk factors to improve long-term health screening for type 2 diabetes and obesity.

**Key messages:** *What is already known on this topic:*

1. Gestational diabetes mellitus (GDM) is associated with maternal and neonatal morbidity, which is substantially reduced with early recognition and management.
2. GDM screening approaches vary substantially in efficiency, diagnostic accuracy, and resource use. Universal screening with a 75g oral glucose tolerance test (75g OGTT) is resource-intensive, yet risk factor-based screening alone risks missing GDM in women without recognised risk factors.
3. The recurrence risk of GDM is up to 41.3%, and women with GDM are more likely to develop type 2 diabetes and cardiovascular disorders; therefore, diagnosis of GDM is important to identify and intervene in those women who are at higher risk.
4. GDM is associated with increased offspring adiposity and childhood obesity, impaired glucose metabolism and a substantially elevated long-term risk of prediabetes and type 2 diabetes, as well as a broader adverse cardiometabolic profile.

*What this study adds:*

1. Universal screening detected GDM in women who may not have been offered screening based on risk factors.
2. Around 1/3 of GDM cases detected in this population were diagnosed later in pregnancy following negative GCT and/or negative OGTT.
3. The relationship between maternal glycaemia and adverse pregnancy outcomes is continuous across GCT glucose levels, with risk increasing with increased glucose levels.

*How this study might affect research, practice or policy:*

1. Further research is needed to investigate how a universal screening approach in diagnosing GDM addresses longer-term cardiometabolic health.
2. Variation in the predictive value of a GCT for GDM by maternal characteristics should be considered when evaluating and optimising any universal screening strategy.
3. A GCT population could form part of a hybrid GDM screening strategy where high-risk women proceed directly to 75g OGTT and women considered at low risk of GDM benefit from screening by GCT.

## Introduction

Gestational diabetes mellitus (GDM) refers to the development of glucose intolerance during pregnancy, arising when placental hormones increase maternal insulin resistance beyond the pancreas’s ability to compensate, leading to hyperglycaemia (1, 2). Nearly one-fifth (19.7%) of live births worldwide are affected by hyperglycaemia, of which around 80% are attributable to GDM rather than pre-existing diabetes (3). Women diagnosed with GDM are at increased risk of developing pre-eclampsia, delivery by caesarean section and, in the longer term, of developing type 2 diabetes mellitus (T2DM) later in life (4). Infants of GDM mothers are at a higher risk of shoulder dystocia, neonatal respiratory distress syndrome, neonatal hypoglycaemia, neonatal jaundice, neonatal unit admission, and also longer-term risk of obesity and early-onset type 2 diabetes (5–7). Consequently, early recognition and management of GDM is vital to optimise perinatal and longer-term outcomes for both mother and offspring.

Screening approaches for GDM vary substantially across different healthcare settings, with longstanding debates about efficiency, diagnostic accuracy and optimal resource use (8). Any screening approach must ultimately do more good than harm across the population, not only for those who benefit from earlier diagnosis, as set out by the United Kingdom (UK) National Screening Committee (9). In the UK, the National Institute for Health and Care Excellence (NICE) supports a one-step, risk factor-based strategy for screening women at risk of developing GDM. In this approach, high risk-group women (i.e., body mass index [BMI] >30, previous macrosomia, previous GDM, first degree relative with GDM, and of high-risk ethnicity, such as South Asian, Black African or Caribbean and Middle Eastern) undergo a single fasting 75g oral glucose tolerance test (75g OGTT) between 24 and 28 weeks’ gestation, with GDM diagnosed if fasting glucose is ≥ 5.6 mmol/L or 2-hour plasma glucose ≥7.8 mmol/L (10). This UK approach is intended to promote an efficient screening pathway to diagnose GDM directly, but relies heavily on identifying women with recognised risk factors for testing and offers no screening to those without those factors (11). Furthermore, women with a negative screening result at the standard gestational age may later be diagnosed with late-onset GDM (defined as after 34 weeks) when clinical features such as glycosuria, polyhydramnios, or fetal macrosomia are detected later in pregnancy (12).

In contrast, countries such as Canada, Australia, and New Zealand have adopted a one-step, universal screening diagnostic approach recommended by the International Association of Diabetes and Pregnancy Study (IADPSG) (13). This involves all pregnant women having a 75 g OGTT at 24-28 weeks, with GDM diagnosed if either fasting glucose ≥ 5.1mmol/L, 1-hour plasma glucose ≥ 10.0mmol/L or 2-hour plasma glucose ≥ 8.5mmol/L. Other countries, such as the United States of America, use a two-step diagnostic pathway, where all pregnant women are invited for a non-fasting 50g glucose challenge test (50g GCT) at 26 to 28 weeks. All women screened positive (1-hour plasma glucose ≥ 7.8mmol/L) are invited for a diagnostic 100g OGTT, with GDM diagnosed when either fasting glucose ≥ 5.1mmol/L, 1-hour plasma glucose ≥ 10.0mmol/L or 2-hour plasma glucose ≥ 8.5mmol/L (14). The 50g GCT being a fasting test for only two hours makes it a convenient initial screening tool, allowing it to be administered easily at any scheduled antenatal appointment, supporting the practicality of universal screening. However, its accuracy varies between populations and may miss women with milder or fasting hyperglycaemia (15).

Given the uncertainty of the 50g GCT as a universal screening tool, there is a need to evaluate its performance and clinical significance among a large and diverse population. This study therefore aimed to assess the maternal and neonatal outcomes among women undergoing universal screening with the 50g GCT, particularly in women who would not have been offered screening by NICE criteria. Further, we assessed the number of women found to have GDM after a negative CGT or positive CGT, but normal OGTT.

## Research design and methods

### Study design and participants

This was a retrospective observational cohort study analysing pregnancies screened for GDM at a large tertiary hospital trust (Chelsea and Westminster Hospital NHS Foundation Trust, London, United Kingdom) between January 2010 and December 2023. All women who had a 50g GCT at 24-28 weeks were included; women with pre-existing type 1 and type 2 diabetes were excluded. During the study period, women with a history of GDM in a previous pregnancy were all offered early capillary blood glucose testing as standard of care and hence excluded from this cohort.

### Clinical pathway

During the study period, the hospital trust followed a two-step universal screening pathway where all women without a history of GDM or pre-existing diabetes had a 50g GCT between 24 and 28 weeks’ gestation. Those with a 50g GCT screening glucose value of ≥ 7.8 mmol/L proceeded to a diagnostic 75g OGTT. GDM was diagnosed if any one of the following thresholds on 75g OGTT were met: fasting glucose ≥ 5.6 mmol/L, 1-hour glucose ≥ 11.0mmol/L, or 2-hour glucose ≥ 7.8 mmol/L.

Women who tested positive for GDM were referred to the Diabetes in Pregnancy clinic, where they received individualised dietary and lifestyle counselling and performed capillary blood glucose monitoring four times daily (target values: fasting <5.3 mmol/L; 1-hour postprandial <7.8 mmol/L). Pharmacological treatment with metformin and/or insulin was initiated when at least 20% of recorded glucose measurements remained above target despite adherence to dietary and lifestyle interventions. Furthermore, some women, despite a normal 75g OGTT at 28 weeks’ gestation, underwent repeat 75g OGTT diagnostic testing later in pregnancy if they developed glycosuria (defined by 1+ on two occasions or 2+ once), polyhydramnios, or if there was a suspicion of a large for gestational age (LGA) baby (estimated fetal weight >95^th^ centile and or abdominal circumference >97^th^ centile) suggestive of late onset GDM (Figure 1).

**Figure 1:**
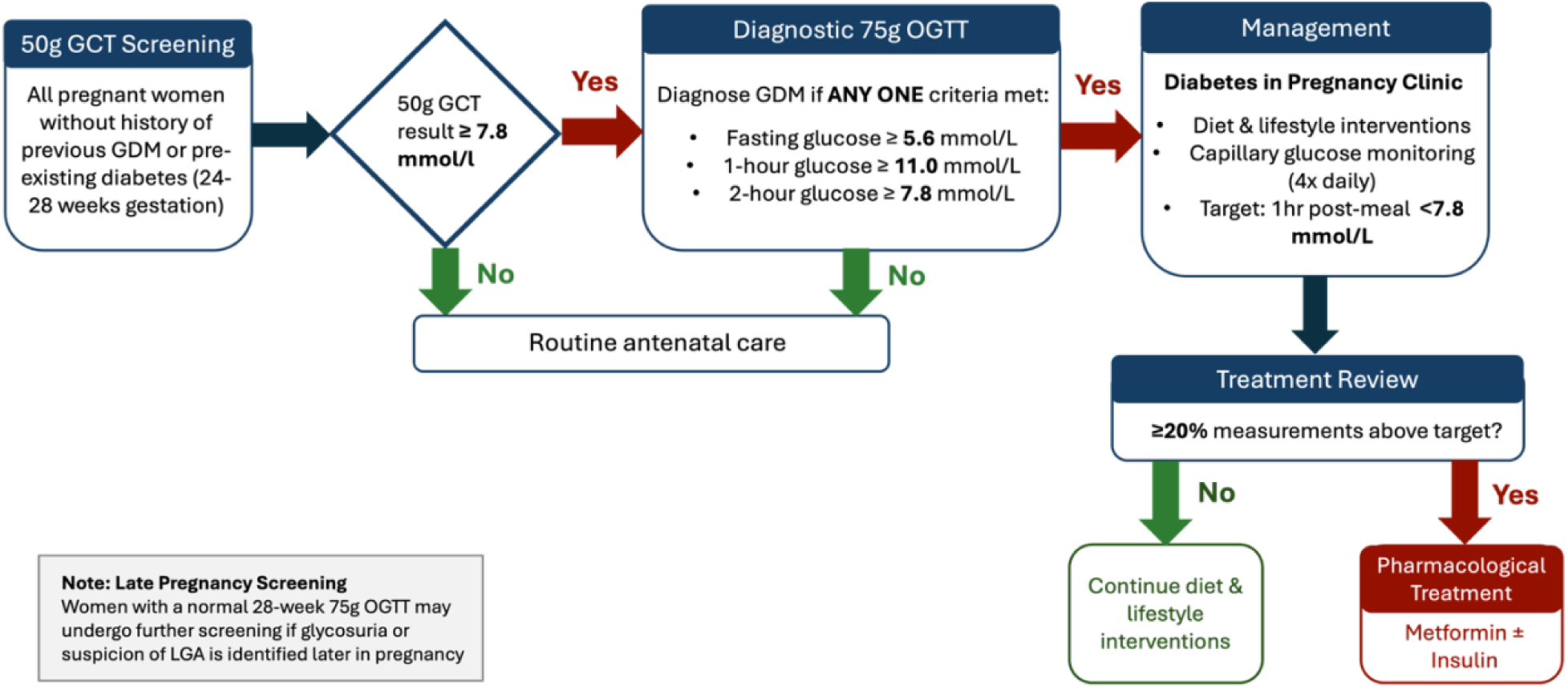
Clinical pathway for GDM Screening and Management. The flow chart illustrates the two-step screening approach between 24 and 28 weeks gestation. All pregnant women (without a history of GDM in a previous pregnancy or pre-existing type 1 and type 2 diabetes) are screened at 24-28 weeks gestation with a 50g GCT. Women with a 50g GCT ≥ 7.8 mmol/L were referred for a diagnostic 75g OGTT, where GDM is diagnosed if any one of the thresholds is met: Fasting: ≥ 5.6 mmol/L, 1-hour ≥ 11 mmol/L, or 2-hour ≥ 7.8 mmol/L. Following diagnosis, patients received lifestyle counselling and initiated capillary glucose monitoring (Targets: fasting less than 5.3mmol/l and 1-hour post-meal <7.8 mmol/L). Pharmacological therapy (metformin and/or insulin) was initiated if ≥ 20% of readings exceeded these targets. Late pregnancy screening was performed for suspected LGA, polyhydramnios or glycosuria regardless of previous 50g GCT or 75g OGTT results.

### Data collection and variables

The Circonia (CMIS), K2 Athena maternity information systems and Cerner Millennium electronic patient records were used to identify women with a singleton pregnancy who had a 50g GCT (n=18,905). We excluded women with multiple pregnancies to reduce bias associated with more adverse maternal and neonatal outcomes (16). 18,213 singleton pregnancies were therefore included in the final analysis. Women were categorised into a screen-negative group (n=16,198), defined by a GCT <7.8mmol/L, and a screen-positive group (n=2,015), defined by a GCT ≥7.8 mmol/L (Figure 2). Furthermore, we identified those women who had a final diagnosis code of GDM to ensure we included those women diagnosed with GDM in the pregnancy at any time point, including outside of the standard screening pathway. Maternal demographics and characteristics were recorded for all groups.

**Figure 2:**
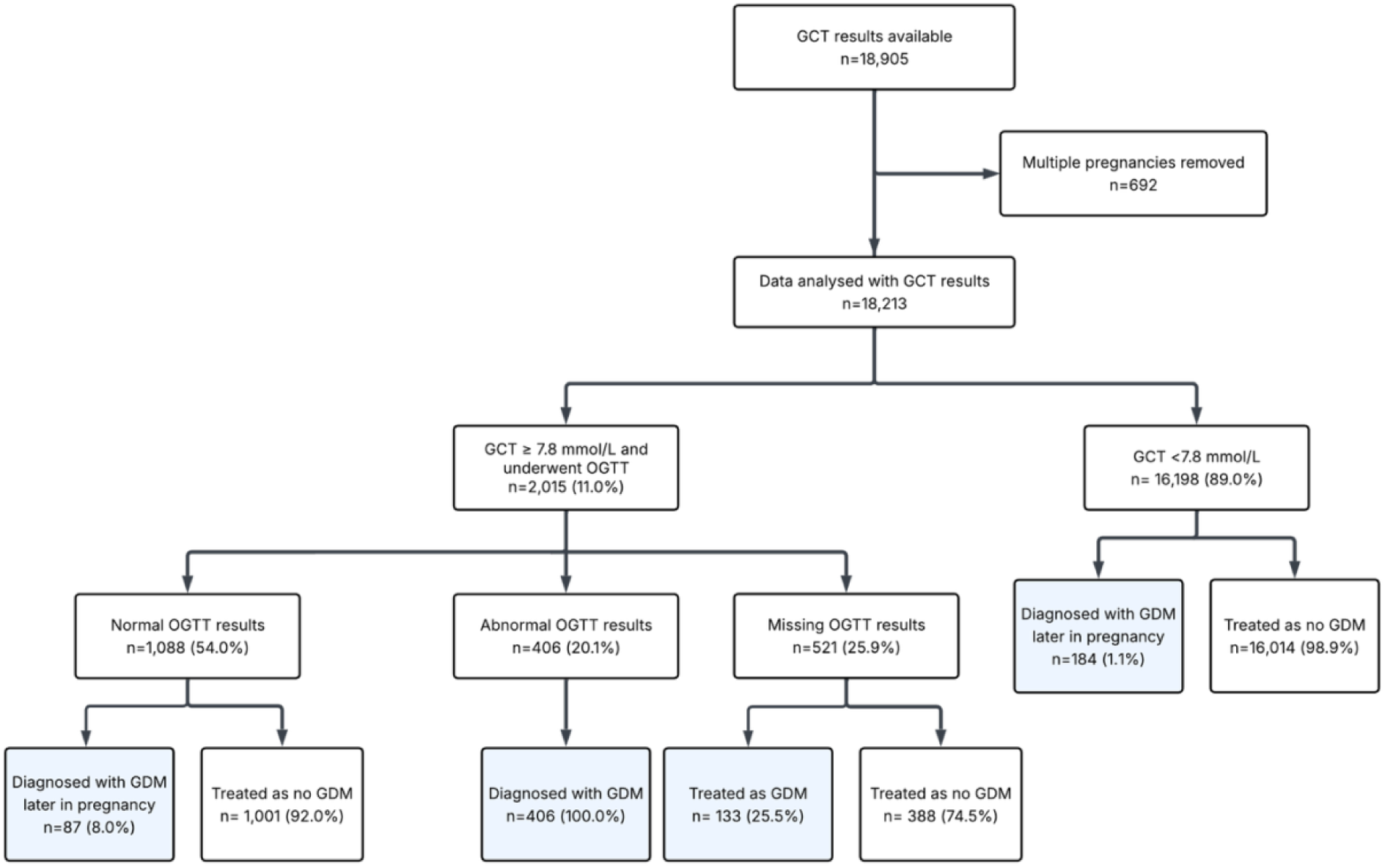
Patient selection and grouping process. Diagnosis of GDM was based on a modified institutional protocol: fasting glucose ≥ 5.6 mmol/L, 1-hour glucose ≥ 11.0mmol/L, or 2-hour glucose ≥ 7.8 mmol/L. Multiple pregnancies, including twins and higher-order multiples were excluded. Missing OGTT is defined as missing one or more components of the 75g OGTT. Abbreviations: GCT, glucose challenge test; GDM, gestational diabetes mellitus; 75g-OGTT, oral glucose tolerance test.

Macrosomia is defined as a birth weight of 4kg of more. Large for gestational age (LGA) is defined as an abdominal circumference >97^th^ centile and/or estimated fetal weight of >90^th^ centile, whereas small for gestational age (SGA) is defined as an estimated fetal weight of <10^th^ centile.

### Statistical analysis

Baseline characteristics and associations between GCT categories and pregnancy outcomes were summarised using a median (interquartile range [IQR]) for continuous variables, and frequencies and percentages for categorical variables. Following normality testing, group comparisons for continuous variables were performed using the Wilcoxon rank-sum or Kruskal-Wallis non-parametric tests (17, 18). Comparisons between groups of categorical data were performed using Fisher’s exact test for expected frequencies <5, and Pearson Chi-squared test for larger counts (19, 20). Statistical significance was defined as a P-value <0.05. Multivariable logistic regression was performed to evaluate the independent association between the screening GCT value and the subsequent diagnosis of GDM. GDM was modelled as a binary dependent variable (GDM diagnosed on a confirmatory 75 g OGTT ‘yes’ versus ‘no’), with screening GCT values entered as a continuous predictor. Effect estimates were expressed as adjusted odds ratios (aORs) with 95% confidence intervals (CIs) per 1.0 mmol/L increase in maternal glycaemia. Models were adjusted for prespecified maternal characteristics associated with GDM based on literature, including age group, BMI group, deprivation quintile and ethnicity (21).

50g GCT values were analysed as a continuous variable without categorisation. To assess the discriminatory ability of 50g GCT for GDM, defined according to the final diagnosis records, receiver operating characteristic (ROC) curves were constructed using 50g GCT as the index test and the final GDM diagnosis as the reference outcome (22). The area under the ROC curve (AUC) and corresponding 95% CIs were estimated using the DeLong method. AUCs were calculated for the overall cohort and within prespecified strata of maternal age, BMI, deprivation index and ethnicity.

Continuous GCT variables were summarised using medians and interquartile ranges (IQRs). Differences across ethnicity, BMI and deprivation groups were assessed using Kruskal-Wallis test, with Dunn’s post-hoc tests and Holm correction. Adjusted linear regression was used to assess associations of ethnicity and BMI with GCT, adjusting for parity and gestational age at screening. A two-sided p < 0.05 was considered statistically significant. All statistical analyses were performed using the R software version 4.2.2 (23).

### Ethical considerations

This study was approved by the Health Research Authority and the local ethics committee (REC reference: 25/HRA/0760; IRAS project ID: 348986). This was a retrospective evaluation of routine care delivery at the hospital Trust, and informed consent of individuals for whom data were analysed was not required. All study procedures were conducted in accordance with institutional guidelines for clinical data use and the principles of the Declaration of Helsinki.

## Results

### Baseline characteristics

Patient demographics and baseline characteristics differed significantly between the GDM and non-GDM cohorts (Table 1). 18,213 women were included in the study, and 810 (4.4%) were diagnosed with GDM. The GDM cohort was older (median 34.0 vs 33.0 years, p<0.001) and gave birth at a significantly earlier median gestational age (38.0 weeks; IQR 38.0-39.0) than the non-GDM cohort (39.0 weeks; IQR 39.0-40.0, p<0.001).

**Table 1:** Patient demographics and baseline characteristics. Data are presented as n (%) or median (IQR). Deprivation levels are presented as quintiles (Q1-Q5) based on the 201S Index of Multiple Deprivation (IMD), where Q1 represents the least deprived and Q5 represents the most deprived.^1^Wilcoxon rank sum test; ^2^Pearson’s Chi-squared test. Abbreviations: GDM, gestational diabetes mellitus; non-GDM, non-gestational diabetes mellitus.

| Characteristic | GDM results (n=18,213) |  | p-value |
| --- | --- | --- | --- |
|  | non-GDM<br>N = 17,403 | GDM<br>N = 810 |  |
| **Gestation At Delivery (Weeks), Median (Q1, Q3)** | 39.00 (39.00, 40.00) | 38.00 (38.00, 39.00) | <0.001<sup>1</sup> |
| **Age, Median (Q1, Q3)** | 33.0 (30.0, 36.0) | 34.0 (31.0, 38.0) | <0.001<sup>1</sup> |
| **Age group, n (%)** |  |  | <0.001<sup>2</sup> |
| ≤ 25 | 1,357 (7.8%) | 49 (6.0%) | 0.068<sup>2</sup> |
| 26 - 30 | 3,668 (21.1%) | 146 (18.0%) | 0.037<sup>2</sup> |
| 31 - 34 | 5,934 (34.1%) | 231 (28.5%) | 0.001<sup>2</sup> |
| > 34 | 6,444 (37.0%) | 384 (47.4%) | <0.001<sup>2</sup> |
| **BMI, Median (Q1, Q3)** | 23.0 (21.0, 25.0) | 25.0 (22.0, 30.0) | <0.001<sup>1</sup> |
| **BMI group, n (%)** |  |  | <0.001<sup>2</sup> |
| < 18.5 | 728 (4.2%) | 16 (2.0%) | 0.002<sup>2</sup> |
| 18.5 - 24.9 | 11,462 (65.9%) | 330 (40.7%) | <0.001<sup>2</sup> |
| 25.0 - 29.9 | 3,690 (21.2%) | 242 (29.9%) | <0.001<sup>2</sup> |
| 30.0 - 34.9 | 1,078 (6.2%) | 138 (17.0%) | <0.001<sup>2</sup> |
| ≥ 35.0 | 445 (2.6%) | 84 (10.4%) | <0.001<sup>2</sup> |
| **Deprivation Decile, Median (Q1, Q3)** | 6.00 (4.00, 8.00) | 5.00 (4.00, 7.00) | <0.001<sup>1</sup> |
| **Deprivation Level, n (%)** |  |  | <0.001<sup>2</sup> |
| Q1 | 1,506 (8.7%) | 87 (10.7%) | 0.040<sup>2</sup> |
| Q2 | 3,897 (22.4%) | 228 (28.1%) | <0.001<sup>2</sup> |
| Q3 | 4,286 (24.6%) | 217 (26.8%) | 0.163<sup>2</sup> |
| Q4 | 4,436 (25.5%) | 174 (21.5%) | 0.010<sup>2</sup> |
| Q5 | 3,278 (18.8%) | 104 (12.8%) | <0.001<sup>2*</sup> |
| **Ethnicity, n (%)** |  |  | <0.001<sup>2</sup> |
| White | 10,125 (58.2%) | 334 (41.2%) | <0.001<sup>2</sup> |
| Asian | 1,903 (10.9%) | 203 (25.1%) | <0.001<sup>2</sup> |
| Black | 1,180 (6.8%) | 73 (9.0%) | 0.014<sup>2</sup> |
| Mixed | 536 (3.1%) | 22 (2.7%) | 0.557<sup>2</sup> |
| Other | 3,659 (21.0%) | 178 (22.0%) | 0.517<sup>2</sup> |

Overall, the GDM cohort exhibited a significantly higher median BMI compared to the non-GDM cohort (25.0 kg/m^2^ vs 23.0 kg/m^2^, p<0.001). The GDM cohort had a lower prevalence of normal-weight individuals (40.7% vs 65.9%, p<0.001) and higher rates of those who were overweight (29.9% vs 21.2%, p<0.001) and obese (27.4% vs 8.8%, p<0.001). Differences in socioeconomic status were also observed, with a lower median deprivation decile (i.e., more deprived) in the GDM group versus non-GDM (5.00 vs 6.00, p<0.001). Ethnic composition also differed significantly, characterised by a lower proportion of White women (41.2% vs 58.2%, p<0.001) and a higher representation of Asian women (25.1% vs 10.9%, p<0.001) within the GDM cohort.

### Diagnosis of GDM

Of the 2,015 women who screened positive with a 50g GCT value of ≥7.8 mmol/L, 75g-OGTT values were available for 1,494 women from the electronic health records, and of these, 406 women had a diagnosis of GDM based on an abnormal 75g-OGTT result, yielding a positive predictive value (PPV) of 27.2%.

### Diagnosis of late-onset GDM

271 women (271/2015, 13.5%) had a diagnosis of late-onset GDM. 184 of these women (184/271, 67.9%) had a negative GCT, and 87 women (87/271, 32.1%) had a positive 50g CGT followed by a negative 75g OGTT.

### GDM diagnosis among women who missed 75g OGTT

Of those with a positive 50g GCT, 521 women (25.9%) did not have a confirmatory 75g OGTT, of which 133 (25.5%) were subsequently diagnosed with GDM later in pregnancy.

### Overall diagnostic accuracy of GCT as a universal screening tool

Among the 18,213 universally screened women, 2,015 (11.1%) had a positive 50g GCT result and 406 women were then diagnosed with GDM with a 75g OGTT, giving a diagnostic yield of 2.23%. The universal screening pathway also detected GDM developed later in the pregnancy following negative or missed screening at 24-28 weeks’ gestation, if additional clinical indications developed. Including later diagnosed cases, the overall diagnostic yield of the universal pathway was 4.4%. ROC analysis was subsequently used to evaluate the discriminatory performance of the GCT result in relation to a final diagnosis of GDM. For the whole universally screened cohort, the ROC analysis demonstrated an area under the curve (AUC) of 0.863 for the discriminatory performance of the GCT for final GDM diagnosis (n = 18,213) (Figure 3). However, this analysis should be interpreted in the context of the screening pathway, since the OGTT was only performed in women with a positive GCT. Definitive OGTT-based GDM status was not available for all women with a negative GCT.

**Figure 3:**
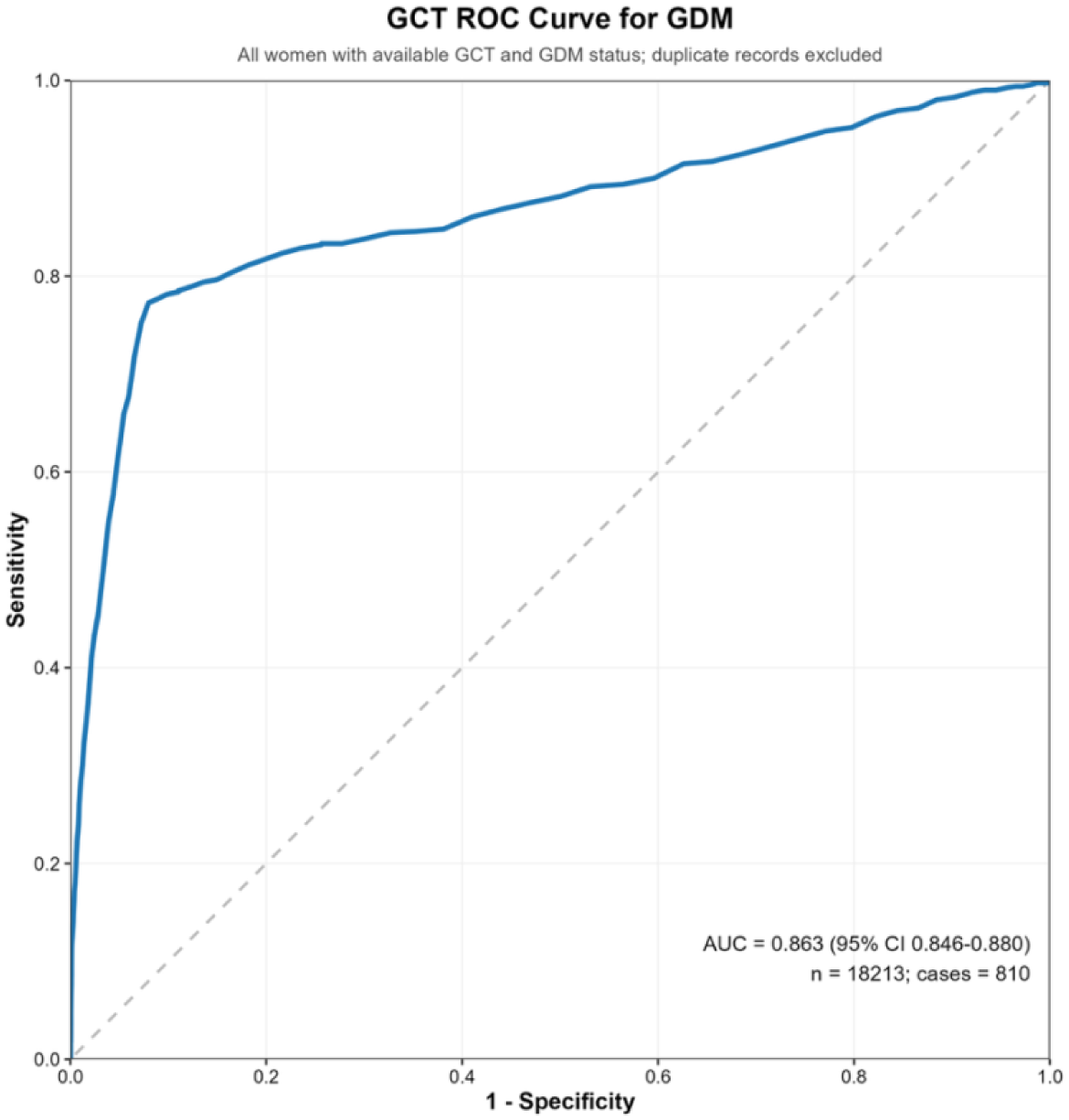
Discriminatory performance of the GCT for GDM in the entire cohort. Receiver Operating Characteristic (ROC) curve displaying the discriminatory performance of the Glucose Challenge Test (GCT) for Gestational Diabetes Mellitus (GDM). The area under the curve (AUC) is 0.8c3 (S5% Cl: 0.84c-0.88). Abbreviations: AUC, Area under the Curve; GCT, Glucose Challenge Test; ROC, Receiver Operating Characteristics.

### Predictive performance of 50g GCT across different maternal subgroups

The predictive performance of 50g GCT for GDM in women was evaluated across subgroups defined by maternal age, BMI, ethnicity and deprivation index by comparing the discriminatory performance of the GCT across thresholds and the PPV at the 7.8 mmol/L threshold.

#### Age

In the age-stratified analyses, the 50g GCT demonstrated the best overall discriminatory performance among women aged 25 years and below, with an AUC of 0.88 (95% CI: 0.83-0.95). Conversely, the discriminatory performance was the lowest among women older than 34 years (AUC:0.85; 95% CI:0.82-0.88). The PPV varied among different age groups, ranging from 27.8% in the 26-30 years group to 34.5% in the ≤ 25 years and >34 years group. (Supplementary Table 1).

#### BMI

In the BMI-stratified analyses, the 50-g GCT demonstrated the best overall discriminatory performance among women with a BMI of <18.5kg/m^2^ (AUC:0.94, 95% CI: 0.87-1.00). Moderate discriminatory performance was observed in the normal BMI (18.5-24.9kg/m^2^) group (AUC:0.87, 95% CI 0.85-0.90), the overweight (25-29.9kg/m^2^) group (AUC 0.85, 95% CI 0.81-0.89) and the class 1 obesity (30.0-34.9kg/m^2^) group (AUC 0.83, 95% CI 0.78-0.87). The lowest performance was seen in the class II obesity (≥35kg/m^2^) group, where the AUC decreased to 0.80 (95% Cl: 0.74-0.86). The PPV of the GCT also varied with maternal BMI, with the lowest in the <18.5 kg/m^2^ BMI group (19.2%) and highest in the ≥35kg/m^2^ BMI group (53.9%). The PPV was 24.8% in the 18.5-24.9 kg/m^2^ BMI group, 34.6% in the 25.0 to 29.9 kg/m^2^ BMI group and 43.2% in the 30-34.9 kg/m^2^ BMI group (Supplementary Table 1).

#### Ethnicity

The 50g GCT demonstrated the highest overall discriminatory performance in the Other ethnicity cohort with an AUC of 0.88 (95% CI 0.86-0.92) and the lowest among Black and Mixed women, with an AUC of 0.81 (95% CI 0.75-0.87) and an AUC of 0.81 (95% CI 0.67 to 0.95), respectively. The discriminatory performance was similar in the White (AUC 0.85, 95% CI 0.83-0.88) and Asian (AUC 0.86, 95% CI 0.83-0.90) subgroups. The PPV across different ethnicity groups ranged from 24.6% in White women to 42.3% in Asian women. (Supplementary Table 1).

#### Deprivation

When stratified by deprivation index, the overall discriminatory performance of the 50g GCT was comparable between quintiles, with an AUC ranging from 0.85 in Q3 and Q4 to 0.88 in Q5. The PPV of the GCT varied with maternal deprivation index, with 36.7% in the Q1 group, 34.3% in the Q2 group, 30.3% in the Q3 group, 29.5% in the Q4 group and 24.1% in the Q5 group (Supplementary Table 1).

### Predictive performance of 50g GCT (≥ 7.8 mmol/L) in low-risk women

The performance of the 50g GCT among the subgroup of women who were at low risk for GDM and would have been excluded from risk factor-based GDM screening (White ethnicity, nulliparous and BMI <30kg/m^2^) was investigated. The incidence of GDM in this group was 2.90% (173/5,972), with an AUC of 0.90 (95% CI 0.86-0.93) for the GCT and a PPV of 24.6% (141/574) at the 7.8 mmol/L threshold.

### Multivariable linear regression comparing glucose variable among different ethnicities, BMI and deprivation index

Given that our results showed variation in discriminatory performance of GCT among subgroups of ethnicity, BMI and age, independent association with each risk factor was explored using adjusted regression modelling. The aim was to determine a relationship with GCT glucose values.

#### Ethnicity

A multivariable linear regression was performed to evaluate the independent association between ethnicity and GCT glucose concentration, after adjusting for maternal age, BMI, parity and gestational age at screening, with ethnicity remaining independently associated with GCT value (p<0.001). The median GCT was highest among Asian women, at 6.1 (5.2-7.3) mmol/L but comparable between the other groups. The median GCT in White women was 5.6 (4.8-6.6) mmol/L, Mixed ethnicity 5.6 (4.7-6.5) mmol/L, Black women 5.7 (4.8-6.6) mmol/L and Other ethnicities (4.9-6.9) mmol/L. When compared with White women, the mean GCT was higher among Asian women (adjusted difference 0.590 mmol/L, 95% CI 0.519-0.660) and the Other ethnicity group (0.171 mmol/L, 0.116-0.226). There were no clear adjusted differences for Black or Mixed women.

#### BMI

Higher BMI was also associated with higher GCT values (p<0.001). The median GCT value showed an increasing trend with increased BMI, with the <18.5 kg/m^2^ group yielding the lowest median GCT value of 5.5 (4.7-6.5) mmol/L and the ≥ 35.0 kg/m^2^ group yielding the highest median GCT value of 6.2 (5.1-7.9) mmol/L. Dunn’s analysis showed significantly higher GCT values in the 25.0-29.9, 30.0-34.9, and ≥35.0 kg/m^2^ groups compared with the <18.5 kg/m^2^ group (p<0.001 for all).

#### Deprivation

GCT values differed significantly across the five deprivation level groups (p<0.001). The median GCT was 5.8 mmol/L in Q1 (4.8-6.8), Q2 (4.9-6.8) and Q3 (4.9-6.8), and decreased to 5.6 mmol/L in both Q4 (4.8-6.6) and Q5 (4.8-6.6). On the analysis using Dunn’s test with Holm correction, Q1 had significantly higher GCT values than Q5 (p=0.035), while Q2 and Q3 had significantly higher GCT values than both Q4 and Q5 (p<0.001 for both). No significant differences were observed between Q1 and Q2, Q1 and Q3, Q2 and Q3, or Q4 and Q5 (Holm-adjusted p≥0.062).

### Overall cohort analysis for maternal and neonatal outcomes

Poor glycaemic control is associated with adverse maternal and neonatal outcomes. Therefore, the relationship between GCT glucose concentration and maternal and neonatal outcomes were examined using logistic regression modelling. GCT glucose concentration was analysed as a continuous variable, with odds ratios (OR) representing the change in odds of each outcome associated with a 1 mmol/L increase in GCT glucose concentration.

When analysed as a continuous variable across the entire study population (n=18,213), the GCT value demonstrated a significant positive association with several maternal and neonatal outcomes. Higher GCT values were significantly associated with a higher risk of preeclampsia (OR 1.12; 95% CI 1.03-1.22; p=0.009) and postpartum haemorrhage (OR 1.24; 95% CI 1.19-1.30; p<0.001) per unit increase in glucose value. Regarding delivery outcomes, higher glucose levels were associated with an increased likelihood of induction of labour (OR 1.10; 95% CI 1.08-1.13; p<0.001), urgent caesarean section (OR 1.16; 95% CI 1.13 – 1.19; p<0.001) and 3rd/4th degree perineal tears (OR 1.17; 95% CI 1.09-1.26; p<0.001) (Figure 4).

**Figure 4.**
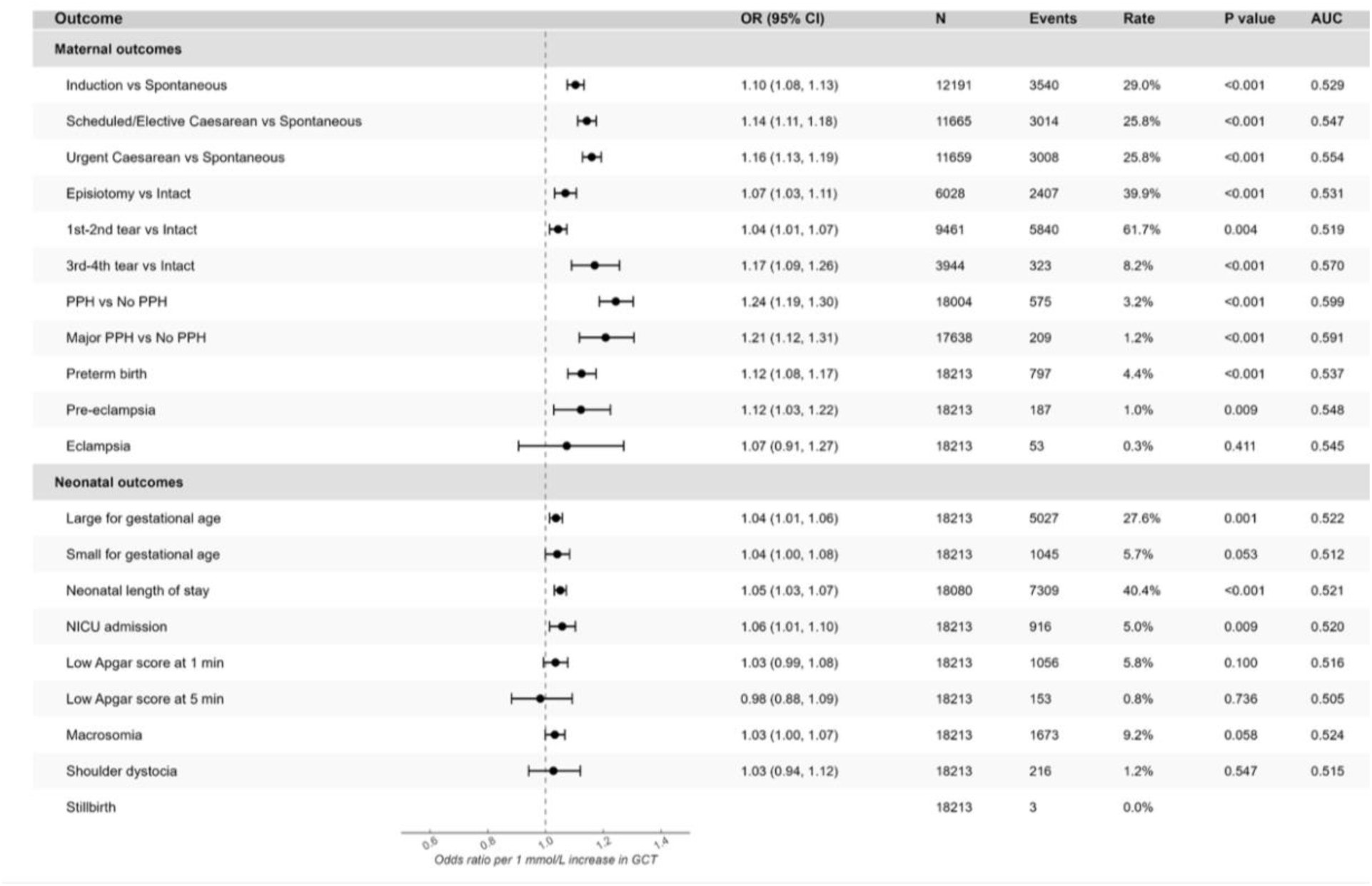
Association of GCT with adverse maternal and neonatal pregnancy outcomes. Forest plot showing odds ratio (OR) and S5% confidence intervals (CI) for the association between GCT result and each outcome, estimated from logistic regression and expressed per 1 mmol/L increase in GCT glucose values. Black dots represent point estimates and horizontal bars the S5% CI; the dashed vertical line marks OR =1 (no association). Outcomes are grouped into maternal and neonatal categories. PPH was defined as a blood loss of more than 500ml within 24 hours of birth, with major PPH defined as a blood loss of more than 1500ml within the same period. Abbreviations: PPH, postpartum haemorrhage; NICU, neonatal intensive care unit.

For neonatal outcomes, significant association was observed between GCT values and preterm birth (OR 1.12; 95% CI 1.08-1.17; p<0.001). Higher glucose levels also significantly increased the odds of LGA infants (OR 1.04; 95% CI 1.01-1.06; p=0.001) and NICU admission (OR 1.06; 95% CI 1.01-1.10; p=0.009). The incidence of stillbirth was low across the entire cohort (n=3), so no meaningful statistical conclusions could be drawn regarding the associations with GCT values (Figure 4).

Among the women considered low-risk for GDM who would have been excluded from risk-factor based GDM screening, the association between certain adverse outcomes (induction of labour, perineal tear, eclampsia, NICU admission, low Apgar score at 1 and 5 minutes, macrosomia and shoulder dystocia) lost statistical significance, but the association with pre-eclampsia (OR 1.15, 95% CI 0.99-1.34), urgent caesarean section (OR 1.06, 95% CI 1.01-1.12), postpartum haemorrhage (OR1.15, 95% CI 1.01-1.31), preterm birth (OR 1.12, 95% CI 1.03-1.21) and LGA (OR 1.04, 95% CI 1.00-1.08) remained (Supplementary Table 2).

## Discussion

### Principal findings

This study provides a large-scale, real-world evaluation of a universal GDM screening pathway utilising the 50g GCT within a diverse UK population (n=18,213), with an observed GDM prevalence and diagnostic yield of 4.4% (810/18213), similar to that from an earlier UK study (11).

In this cohort, the GCT identified 77.3% of women who were diagnosed with GDM. Interestingly, almost 33.5% (271/810) of GDM cases were diagnosed as late-onset GDM despite having negative screening for GDM earlier in gestation, highlighting the limitations of a single screening assessment for a condition that may develop progressively through pregnancy.

#### Implication for risk factor-based screening

Current UK NICE guidelines advocate for a selective screening strategy for GDM based on predefined maternal risk factors, BMI over 30, previous GDM, family history of diabetes and high-risk ethnicity (10). Women considered low risk under NICE screening criteria, who would not be offered GDM screening under current guidance, accounted for nearly one-third of the total study cohort (5,972/18,213) and had a 2.9% prevalence of GDM. Similar to Meek et al, we found that within this low-risk subgroup, elevated GCT values were significantly associated with adverse clinical outcomes, including labour type (scheduled/ elective caesarean vs spontaneous and urgent caesarean vs spontaneous), preterm birth, LGA infants, and perineal tear (3^rd^ and 4^th^ degree tear vs intact perineum) (11). These findings suggest a risk-factor-based approach to GDM screening misses a sub-group of women who, despite the absence of GDM risk factors, have clinically relevant dysglycaemia and are vulnerable to adverse pregnancy outcomes. These findings highlight a central trade-off between efficiency and equity in GDM screening policy. Universal GCT screening offers a potential advantage over selective risk factor-based screening by improving case ascertainment and equity of access, at the cost of increased initial testing.

Arguments against universal screening focus on overdiagnosis and its cost implications (24). Given the alarming increases and growing burden of early-onset type 2 diabetes, especially in the UK population, it may be time to rethink the balance between risks and benefits of universal screening and detection (25). The joint statement by FIGO, EBCOG and EAPM, together with the Barcelona declaration, has acknowledged that a universal screening approach is urgently needed to address the clinical disparity that exists with risk factor-based screening (26, 27). New approaches should be explored to provide screening to women without traditional risk factors. For example, it may be possible to implement a hybrid screening strategy whereby women with established GDM risk factors proceed directly to diagnostic 75g OGTT, while those without GDM risk factors undergo 50g GCT screening and this approach may reduce the need for OGTT by 50% (28).

Failure to complete recommended screening may also have adverse perinatal consequences among women with established GDM risk factors. In this study, 25.9% of those with a positive GCT missed confirmatory OGTT. In a population-based cohort study, non-adherence to GDM screening was associated with a 2.5-fold higher risk of stillbirth among women with pre-existing GDM risk factors, highlighting the potential clinical consequences of missed screening in this group (29). Further research is needed to compare women’s preferences for, and uptake of, the OGTT when it is offered based on risks factors versus a positive GCT.

#### Intersectional performance of 50g GCT as a screening test

Like others, we found that in this UK population, women with GDM were older, with higher BMI, more likely to be of Asian ethnicity and from more deprived areas (30). If the study had adopted a screening strategy based solely on NICE risk-based criteria, this would have disproportionately excluded women, such as those in older age groups without other risk factors from screening, potentially contributing to inequities in GDM detection and diagnosis. Given there is now a body of emerging evidence that GDM represents an early clinical signal of a long-standing trajectory of cardiometabolic risk, this further strengthens the evidence for the need to identify women at increased risk before the onset of overt type 2 diabetes (31).

We further investigated whether there is variation in the predictive performance of the 50g GCT for population subgroups defined by age, BMI, ethnicity and deprivation level. The study found the predictive performance of the GCT at the 7.8 mmol/L threshold does differ substantially between population subgroups, with particularly marked variation by maternal BMI and ethnicity. These subgroup differences were further supported by the multivariable regression analysis, which showed that ethnicity and BMI remained independently associated with GCT after adjustment for parity and gestational age at screening. This may be reflective of the biological heterogeneity in glucose metabolism and GDM aetiology, with differing relative contributions of insulin resistance and insulin secretory defects (32). A large US population-based study also found the glucose threshold values showed ethnic variation, with the highest glucose levels in Filipinos and Chinese, while African American and Caucasian groups had the lowest glucose (30). Collectively, these findings highlight the importance of considering population heterogeneity when evaluating GDM screening strategies, suggesting a single screening threshold may not perform equally well across diverse populations.

#### Strengths and limitations

The strengths of this study are its large cohort size (n=18,213) and population diversity, which enhances the external validity of these findings within the diverse UK population and addresses a limitation in previous studies of the performance of GCT screening. However, definitive characterisation of GCT performance across the full glycaemic spectrum would require a large-scale, multicentre prospective study in which both 50g GCT and confirmatory 75 g OGTT are performed in all participants, regardless of initial screening result.

## Conclusion

Universal screening with the 50 g GCT is a safe and achievable approach in a diverse UK population. It offers an equitable strategy for improving GDM detection across demographic sub-groups, thereby reducing missed diagnosis. Further research is needed to determine how maternal characteristics influence cut-offs for the universal 50g GCT to allow for precision in screening and diagnosis.

## Data Availability

Participant data held locally and cannot be uploaded in full.

## Acknowledgements

The study team recognises the contribution of Honghong Huang and Wei Liu, who initially investigated this topic at the study centre as part of their Master’s projects at Imperial College London.

## Supplementary tables

**Supplementary table 1:**
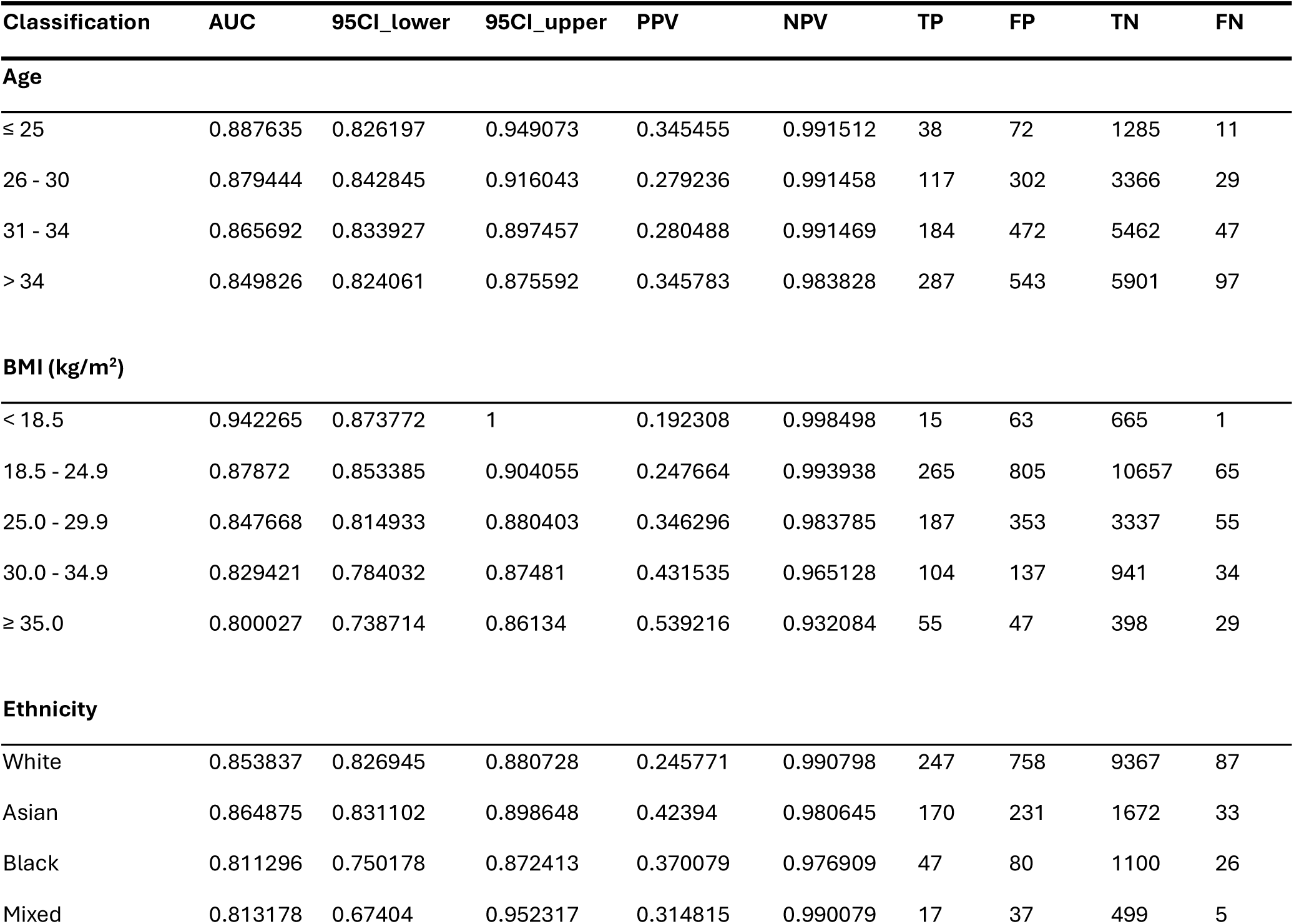
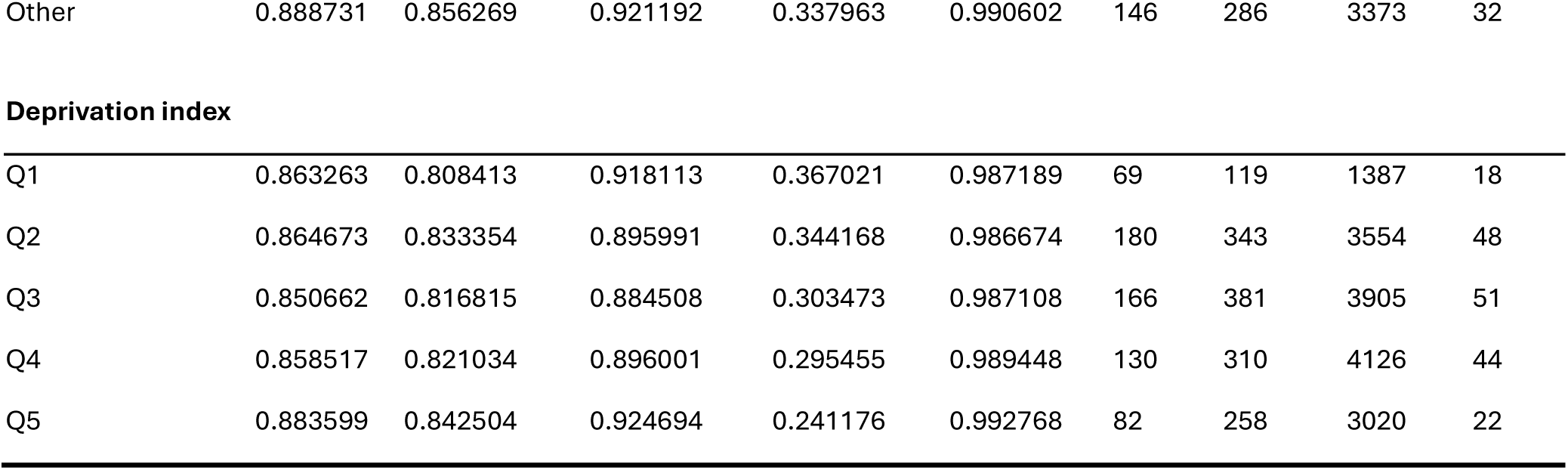
Predictive performance of GCT stratified by age, BMI, ethnicity and deprivation index. Predictive performance of GCT stratified by age, BMI, ethnicity and deprivation index. Abbreviations: AUC = area under the curve; S5 CI_lower/upper = 95% confidence interval bounds; Sens = sensitivity; Spec = specificity; PPV = positive predictive value; NPV = negative predictive value; TP/FP = true/ false positives; TN/FN = true/false negatives; n= sample size.

| Classification | AUC | 95CI_lower | 95CI_upper | PPV | NPV | TP | FP | TN | FN |
| --- | --- | --- | --- | --- | --- | --- | --- | --- | --- |
| **Age** |  |  |  |  |  |  |  |  |  |
| ≤ 25 | 0.887635 | 0.826197 | 0.949073 | 0.345455 | 0.991512 | 38 | 72 | 1285 | 11 |
| 26 - 30 | 0.879444 | 0.842845 | 0.916043 | 0.279236 | 0.991458 | 117 | 302 | 3366 | 29 |
| 31 - 34 | 0.865692 | 0.833927 | 0.897457 | 0.280488 | 0.991469 | 184 | 472 | 5462 | 47 |
| > 34 | 0.849826 | 0.824061 | 0.875592 | 0.345783 | 0.983828 | 287 | 543 | 5901 | 97 |
| **BMI (kg/m<sup>2</sup>)** |  |  |  |  |  |  |  |  |  |
| < 18.5 | 0.942265 | 0.873772 | 1 | 0.192308 | 0.998498 | 15 | 63 | 665 | 1 |
| 18.5 - 24.9 | 0.87872 | 0.853385 | 0.904055 | 0.247664 | 0.993938 | 265 | 805 | 10657 | 65 |
| 25.0 - 29.9 | 0.847668 | 0.814933 | 0.880403 | 0.346296 | 0.983785 | 187 | 353 | 3337 | 55 |
| 30.0 - 34.9 | 0.829421 | 0.784032 | 0.87481 | 0.431535 | 0.965128 | 104 | 137 | 941 | 34 |
| ≥ 35.0 | 0.800027 | 0.738714 | 0.86134 | 0.539216 | 0.932084 | 55 | 47 | 398 | 29 |
| **Ethnicity** |  |  |  |  |  |  |  |  |  |
| White | 0.853837 | 0.826945 | 0.880728 | 0.245771 | 0.990798 | 247 | 758 | 9367 | 87 |
| Asian | 0.864875 | 0.831102 | 0.898648 | 0.42394 | 0.980645 | 170 | 231 | 1672 | 33 |
| Black | 0.811296 | 0.750178 | 0.872413 | 0.370079 | 0.976909 | 47 | 80 | 1100 | 26 |
| Mixed | 0.813178 | 0.67404 | 0.952317 | 0.314815 | 0.990079 | 17 | 37 | 499 | 5 |
| Other | 0.888731 | 0.856269 | 0.921192 | 0.337963 | 0.990602 | 146 | 286 | 3373 | 32 |

*Supplementary table 1: Predictive performance of GCT stratified by age, BMI, ethnicity and deprivation index.
|  |  |  |  |  |  |  |  |  |  |
| --- | --- | --- | --- | --- | --- | --- | --- | --- | --- |
| Q1 | 0.863263 | 0.808413 | 0.918113 | 0.367021 | 0.987189 | 69 | 119 | 1387 | 18 |
| Q2 | 0.864673 | 0.833354 | 0.895991 | 0.344168 | 0.986674 | 180 | 343 | 3554 | 48 |
| Q3 | 0.850662 | 0.816815 | 0.884508 | 0.303473 | 0.987108 | 166 | 381 | 3905 | 51 |
| Q4 | 0.858517 | 0.821034 | 0.896001 | 0.295455 | 0.989448 | 130 | 310 | 4126 | 44 |
| Q5 | 0.883599 | 0.842504 | 0.924694 | 0.241176 | 0.992768 | 82 | 258 | 3020 | 22 |

**Supplementary table 2:**
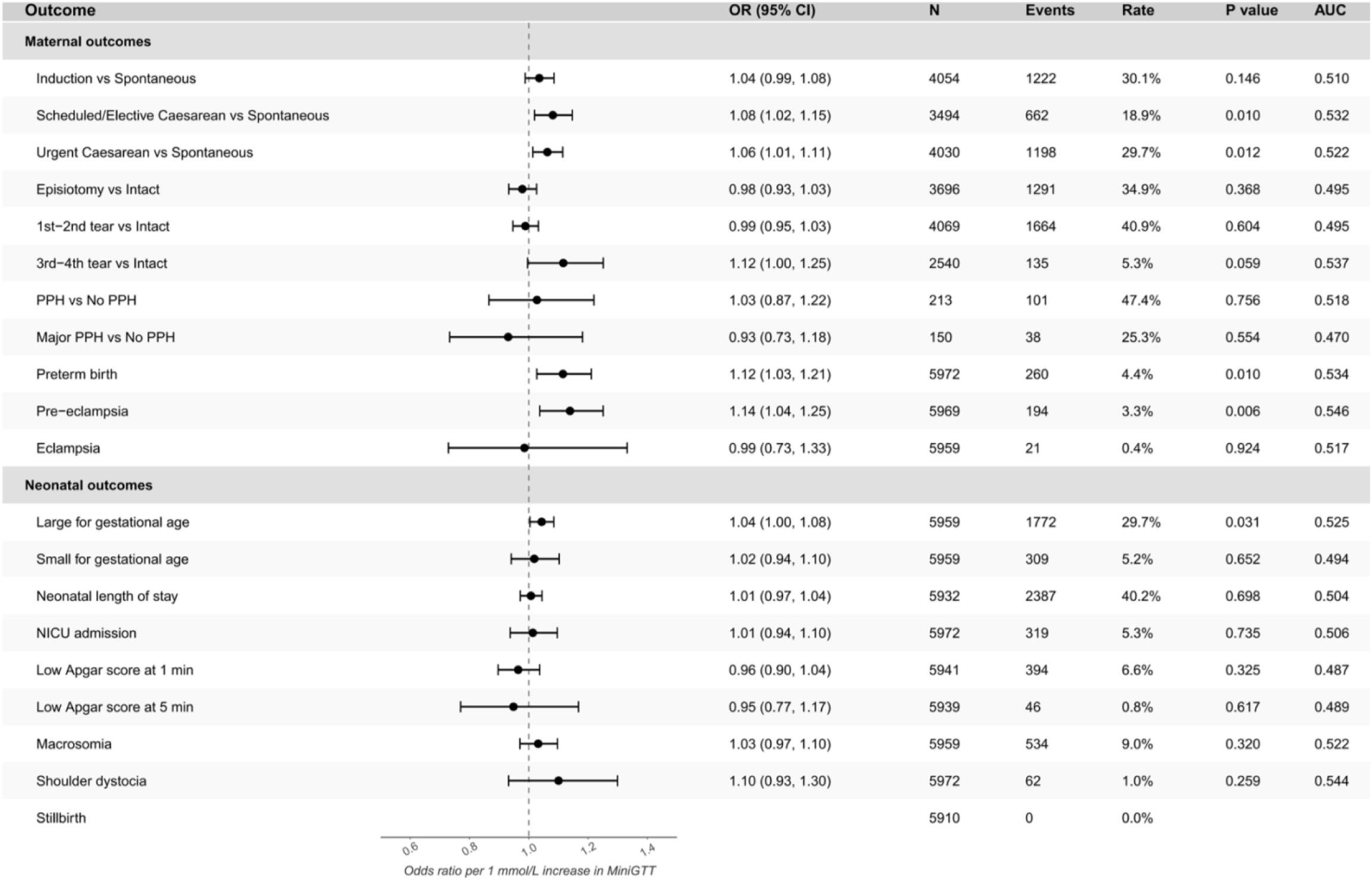
Association between GCT and adverse pregnancy outcomes in women without risk factors for GDM. This forest plot shows the association between GCT and each maternal or neonatal outcome for the low-risk cohort, expressed as the odds ratio for every 1 mmol/L increase in GCT. Each row represents a separate binary comparison, with the black dot showing the estimated odds ratio and the horizontal line showing the 95% confidence interval. Stillbirth was excluded from the logistic regression analysis due to zero events. N represents the number of records included in that specific outcome analysis after excluding missing values; events represent the number of women within N who had the target outcome, coded as 1. The rate is calculated as Events divided by N. Abbreviations: OR, odds ratio; Cl, confidence interval; AUC, area under the receiver operating characteristic curve; PPH, postpartum haemorrhage; NICU, neonatal intensive care unit.

